# Large-Scale EHR-Based Phenotyping of an *FMR1* Premutation Cohort at a Referral Hospital

**DOI:** 10.64898/2026.09.07.26362452

**Authors:** Aadil Rasheed, Jonas Ebner, Akhyar Ahmed, Lothar H. Wieler, Girish N. Nadkarni, Reymundo Lozano, Esther-Maria Antao

## Abstract

**Background:** Rare genetic diseases collectively affect hundreds of millions of people, yet their clinical features are usually defined in small and selected patient groups which do not represent the entire population. The *FMR1* CGG repeat expansion, one of the most common trinucleotide disorders, illustrates what we know about a genetic carrier’s susceptibility that predisposes to multiple medical problems, which unfortunately remains poorly characterised. *FMR1* premutation-associated conditions (FXPAC) encompass a spectrum of clinical manifestations. *FMR1* premutation carriers are at risk of developing conditions such as fragile X–associated primary ovarian insufficiency (FXPOI), fragile X–associated tremor/ataxia syndrome (FXTAS), and a range of neuropsychiatric symptoms collectively termed Fragile X-associated neuropsychiatric disorders (FXAND), which to this date remain a topic of concern and debate. Despite an estimated carrier prevalence of 1 in 151 females and 1 in 468 males, *FMR1* premutation carriers frequently remain undiagnosed or misdiagnosed. Electronic Health Records (EHRs) structured in the Observational Medical Outcomes Partnership (OMOP) Common Data Model (CDM) enable large-scale retrospective phenotyping of such rare conditions.

**Methodology:** Using OMOP-mapped EHR data from the Mount Sinai Health System, we identified a cohort of 750 individuals with a confirmed *FMR1* premutation status (98.5% female; median age at index 33.3 years), to our knowledge the largest EHR-derived *FMR1* premutation cohort described within a single health system to date.

**Results:** Several key FXPAC-associated diagnoses were documented before formal *FMR1* premutation identification: Endocrine/Metabolic (16.5%, n = 124/750) and Neuropsychiatric (16.3%, n = 122/750) conditions were the most prevalent FXPAC domains. In every domain a substantial share of patients had their first record before the index date: the first-quartile lead time was 1.9 years for FXAND (n = 122), 1.5 years for FXPOI (n = 64), 1.0 years for both pain/fatigue (n = 19) and autoimmune/inflammatory conditions (n = 15), and 0.1 years for endocrine/metabolic conditions (n = 124); only FXTAS (n = 13) showed no pre-index lead (Q1 = 0.0 years). These lags reveal a measurable diagnostic gap prior to the identification of the *FMR1* premutation. This study demonstrates that large-scale EHR-based phenotyping can characterise the clinical burden of rare genetic conditions, quantify diagnostic delays at the population scale, and identify precursor diagnosis patterns that may support earlier genetic referral in clinical practice.

## 1. Introduction

The phenotypic features of human genetic disease are typically delineated in selected groups of patients whose ascertainment is biased toward specific subpopulations; these groups rarely represent the full population that carries the genetic variant. Ascertainment bias can propagate through research and into the health care of millions of people worldwide ^1^. Digital health and artificial intelligence are beginning to change this paradigm. Applied to the most common genetic conditions, these technologies offer a real opportunity to characterize diseases as they occur in the general population rather than as they present to medical specialists and after the patients meet diagnostic criteria. In this study, we focus on the *FMR1* premutation, one of the most common nucleotide repeat expansion disorders, with an estimated prevalence of approximately 1 in 151 females and 1 in 468 males^2^. Although recent studies have increasingly sought to clarify which conditions are associated with the *FMR1* premutation ^1,3–11^, the strength and specificity of these associations remain a matter of concern and debate. The *FMR1* premutation, therefore, serves both as a population to be studied and as a test case for how genetic disorders can be characterised using technological advances in digital health and artificial intelligence.

The fragile X premutation-associated conditions (FXPAC) are part of the spectrum of fragile X-related disorders caused by CGG-repeat expansions in the *FMR1* gene. Individuals with 55 to 200 CGG repeats are considered to have an *FMR1* premutation, whereas expansions exceeding 200 CGG repeats are defined as a full mutation and are typically associated with methylation and silencing of *FMR1*. Individuals with a full mutation present with a range of neurodevelopmental conditions, including neurological and behavioural disorders, cognitive and sensory problems, intellectual disability, and autism ^12–14^. Premutation carriers, in contrast, are at increased risk for a distinct set of disorders, including fragile X-associated primary ovarian insufficiency (FXPOI), fragile X-associated tremor/ataxia syndrome (FXTAS), and fragile X-associated neuropsychiatric disorders (FXAND), which together are classified as FXPAC ^15,16^. While the clinical manifestations of the full mutation are well characterised, the phenotypic spectrum among premutation carriers remains incompletely understood. The full spectrum and prevalence of clinical manifestations of the *FMR1* premutation, the determinants of progression to FXTAS or FXPOI, and the variability of neuropsychiatric, cognitive, and other medical problems remain topics of debate.

Most *FMR1* premutation carriers described in the literature were tested only after a relative was diagnosed with fragile X syndrome. Findings derived from such samples may overestimate, misattribute, or, in some domains, obscure true associations ^1,7^. With growing awareness of the prevalence and impact of the premutation, *FMR1* carrier testing is now offered as part of a routine prenatal genetic screening in the United States, allowing pregnant individuals to be tested and families at risk to be identified early. As a result, the Mount Sinai Health System has accumulated a substantial cohort of *FMR1* premutation carriers identified largely through screening rather than through clinical presentation. Launched in 2020, the AI-Ready Mount Sinai (AIR·MS) platform supports large-scale AI-driven clinical research ^17^. The platform integrates data across the Mount Sinai Health System and provides HIPAA-compliant access to data and analytic tools. AIR·MS includes EHR data from more than 12 million patients, encompassing clinical notes, laboratory results, pathology data, and radiologic imaging. The availability of detailed longitudinal EHR data from a cohort of *FMR1* premutation carriers provides a valuable resource for comprehensive clinical characterisation and large-scale phenotypic analyses.

In this study, we retrospectively phenotype a cohort of *FMR1* premutation carriers using AIR·MS, and we characterise the clinical conditions in this population. Specifically, we describe the prevalence of neurological, psychiatric, reproductive, endocrine, and metabolic disorders. We also examine the temporal relationship between clinical diagnosis and identification as an *FMR1* premutation carrier. This dataset likely represents one of the largest cohorts of *FMR1* premutation carriers to be phenotypically described, and our findings have direct implications for earlier detection and better clinical management.

## 2. Methods

### 2.1 Study Design and Data Source

This was a retrospective, observational cohort study of individuals with a documented *FMR1* premutation identified from the electronic health records (EHRs) of the Mount Sinai Health System (MSHS), New York, NY, USA. Data were accessed through the AI-Ready Mount Sinai (AIR·MS) platform, a multimodal health data resource launched in 2020 that provides unified access to EHR, omics, imaging, and sensor data across the health system^17^. AIR·MS holds data for more than 12 million patients, including structured EHR fields, clinical notes, and metadata from pathology and radiology. The study was approved by the Icahn School of Medicine at Mount Sinai Institutional Review Board [STUDY-19-00951: HPIMS Data Science Protocol] with a waiver of informed consent for secondary use of de-identified clinical data.

### 2.2 Source Data Standardisation

Source EHR systems, such as Epic, collect data during the delivery of healthcare services. However, these data are typically stored in proprietary formats, limiting their reuse and interoperability. To address this, the data undergo an extract, transform, and load (ETL) process and are mapped to the Observational Health Data Sciences and Informatics (OHDSI) OMOP CDM using OHDSI standardised vocabularies. AIR·MS extracts data from clinical sites across the Mount Sinai Health System, transforms them into the standardised OMOP CDM format, and loads them into SAP HANA, a high-performance in-memory database. This provides an efficient interface for querying the full OMOP-formatted EHR cohort using Structured Query Language (SQL).

We identified the study cohort using the AIR·MS platform by executing SQL queries against the OMOP CDM and applying standard OMOP concepts. Defining eligibility criteria with standard concepts, rather than raw Epic master-file codes, enabled a single criterion to capture clinically equivalent records across source coding systems and time periods. This approach also supports reuse of the cohort definition at other institutions that use the OMOP CDM. Queries primarily drew on the PERSON, CONDITION_OCCURRENCE, and MEASUREMENT domains.

### Case ascertainment and cohort definition

Most of the patients in our cohort are expected to be young adult females who were recommended *FMR1* genetic testing during prenatal care as part of the Mount Sinai Reproductive Genetic Counselling Program. While screening tests for the *FMR1* premutation and *FMR1* full mutation are well represented as orders in the measurement and procedure domains, the corresponding results were available in a range of formats. Many records referenced a free-text comment, external note, or PDF, and some captured administrative information (e.g., test not performed, cancelled, insufficient quantity, or sent out). A smaller proportion included a machine-readable CGG repeat count or result band.

Classification of *FMR1* premutation (55–200 CGG repeats) versus full mutation (>200 CGG repeats) based on genetic test results is therefore not feasible at scale, and the populations identified through testing and diagnosis show limited overlap. Hence, cohort membership was defined from the recorded code assigned by healthcare providers rather than from *FMR1* genetic test results. *FMR1* premutation carrier status was identified from patients based on fragile X-specific evidence, comprising fragile X carrier status (standard OMOP concept 37109485)

### Pregnancy ascertainment

As most *FMR1* premutation carriers at Mount Sinai are expected to be identified through prenatal genetic screening, we first identified the pregnant population in the health records by integrating information across multiple OMOP tables. A patient was classified as having pregnancy evidence only if a pregnancy-related diagnosis code (condition_occurrence) was accompanied by a positive pregnancy laboratory result (measurement; serum or urine hCG), or by a delivery or pregnancy-loss procedure record (procedure_occurrence). Patients with a diagnosis code alone were not counted. Each patient was counted once irrespective of the number of pregnancy episodes, and the proportion was calculated among females in the cohort (n = 739).

### Index date and longitudinal analysis

The index date was defined as the date of the first recorded fragile X-related diagnosis, taken as the earliest across diagnosis dates. This is the date of first documentation within the health system and is not equivalent to the date of clinical diagnosis or the date of biological disease onset, neither of which can be ascertained in observational EHR data. Longitudinal analyses were conducted relative to the index date, with each patient’s record partitioned into pre- and post-index periods. Comorbidity occurrences and clinical event counts were assigned to the period in which they fell, with same-day events recorded separately. The time from index to first comorbidity occurrence was summarised as a median signed interval in years, negative values indicating occurrence before index. Because the pre-index window is bounded by the start of available records, whereas the post-index window extends to the extraction date, the two periods are unequal in length and vary across patients. Comparisons of counts between periods therefore reflect differential observation time in addition to any change in clinical activity and are reported descriptively rather than as rates.

### Comorbidity ascertainment

Comorbidities were ascertained using a study-specific taxonomy mapping of disease codes to seven categories: neuropsychiatric, neurologic, reproductive and ovarian, endocrine and metabolic, pain and fatigue, autoimmune and inflammatory, and other conditions. Categories were specified *a priori* to reflect the recognised phenotypic spectrum associated with the *FMR1* premutation, including fragile X-associated neuropsychiatric disorders, tremor/ataxia syndrome and primary ovarian insufficiency. The taxonomy is maintained as a single source of truth in the analysis code and is applied identically across all strata.

### Healthcare utilisation

Healthcare utilisation was quantified as per-patient counts of conditions, procedures, measurements, drug exposures, and visits, computed separately for the pre-index and post-index periods. Counts are summarised as medians with interquartile ranges; distributions are strongly right-skewed, and means are reported only where the skew is itself of interest. These measures serve as a proxy for the density of a patient’s engagement with the health system, and hence for the opportunity available for any given condition to be recorded. They are used in this study to assess whether observed differences in comorbidity prevalence between strata are attributable to differential ascertainment rather than to differential disease burden.

### Cohort verification and chart review

A portion of the *FMR1*-Premutation cohort identified programmatically from AIR·MS was subsequently verified through manual patient chart review to confirm the accuracy of cohort identification.

## Results

### Cohort identification

*FMR1* premutation carriers were identified using the Fragile X–specific carrier concept (37109485). Among *FMR1* premutation carriers, we excluded 36 individuals with a diagnosis of *FMR1* full mutation, leaving 750 patients with a confirmed diagnosis (Figure 1). All subsequent analyses were conducted using the resulting cohorts.

**Figure 1:**
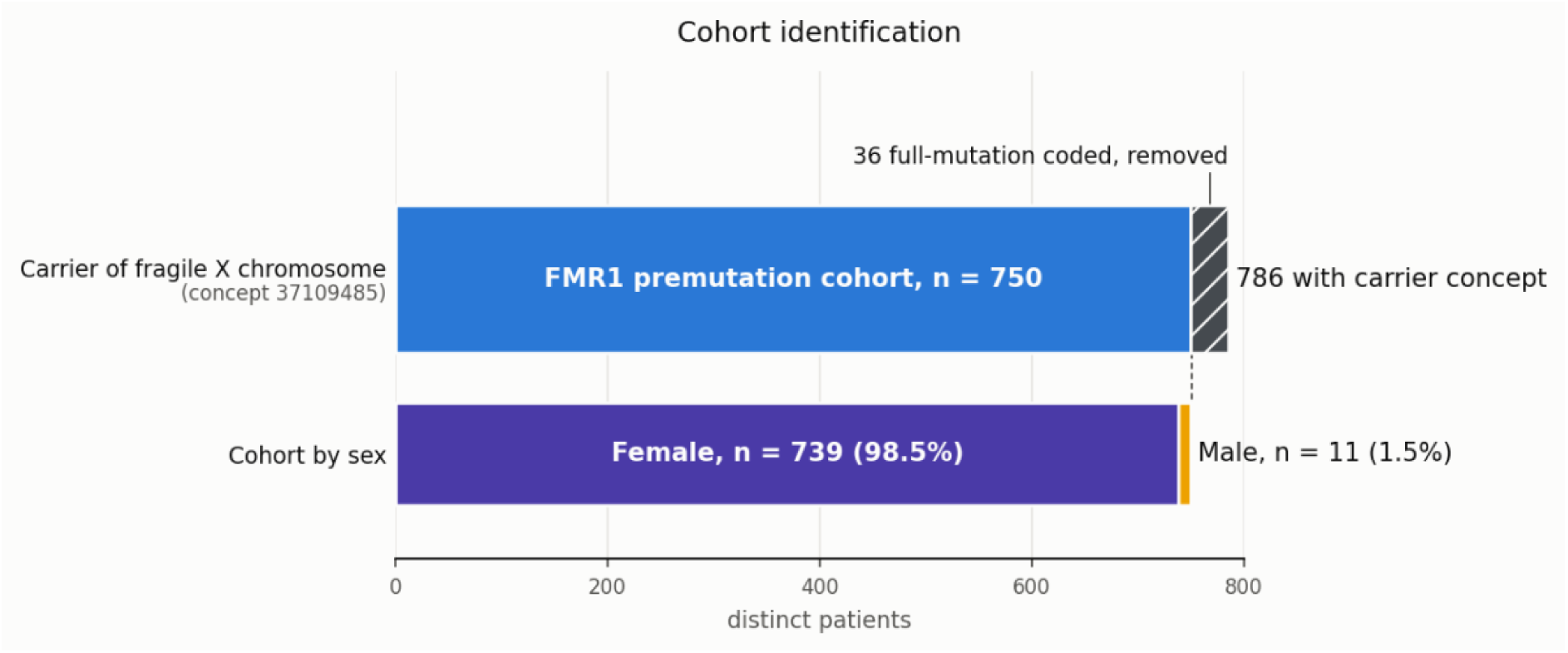
Refinement of the confirmed *FMR1* premutation cohort. Among 786 patients identified using Fragile X-specific diagnostic concepts, 36 individuals with a co-occurring diagnosis of Fragile X syndrome (full-mutation) were excluded, yielding a final cohort of 750 *FMR1* premutation carriers (739 female, 11 male) whose EHRs were used in all subsequent analyses.

### Age-Stratified Clinical Profiling

The confirmed *FMR1* premutation cohort was predominantly composed of women aged 26–45 years (Figure 2). This pattern is consistent with identification through prenatal or reproductive carrier-screening programs at MSHS. We also see that most male identification is distributed across the lifespan (Figure 2).

**Figure 2:**
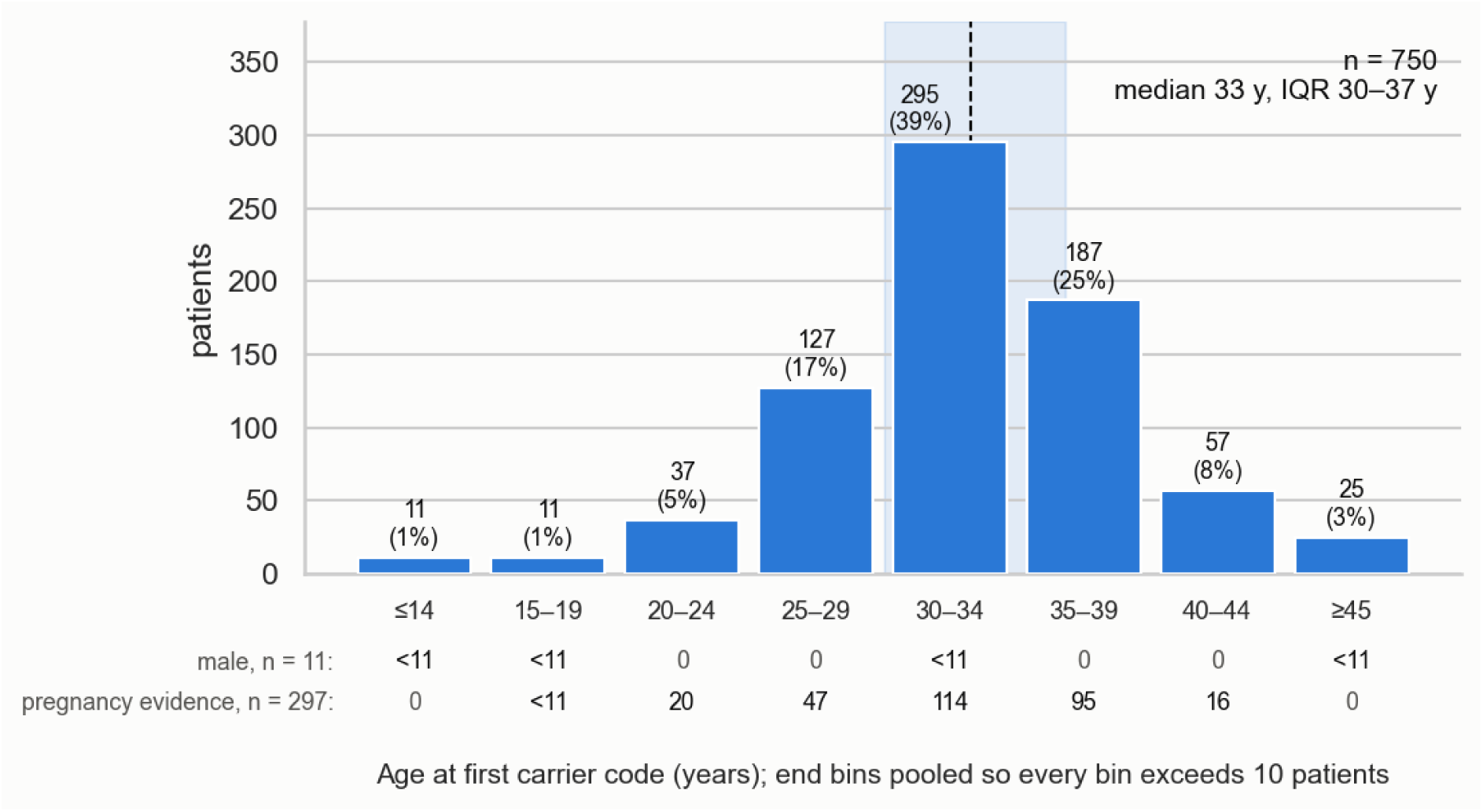
Age at first recorded *FMR1* premutation carrier code in the confirmed cohort (n = 750). Histogram of patient counts in 5-year age bins; end bins are pooled so that every bin exceeds 10 patients. Labels give n (% of cohort). The dashed line marks the median (33 years; IQR 30–37), with the interquartile range shaded. The rows beneath the axis give, per bin, the number of male patients (n = 11) and the number of patients with documented pregnancy evidence (n = 297); counts of 1–10 are shown as <11.

### Demographic structure

The confirmed *FMR1*-premutation cohort was predominantly female (739 of 750 patients; 98.5%). Most patients (87.3%) were aged 26 to 45 years (Figure 2). The median age at index was 33.3 years (IQR, 30.0–36.8). Among females in the confirmed *FMR1* premutation cohort, 297 of 739 (40.2%) had laboratory-confirmed pregnancy evidence (Figure 3). Together, the predominance of women of reproductive age, narrow age distribution, and high prevalence of pregnancy episodes are consistent with reproductive carrier screening mitigated identification (Figure 4).

**Figure 3:**
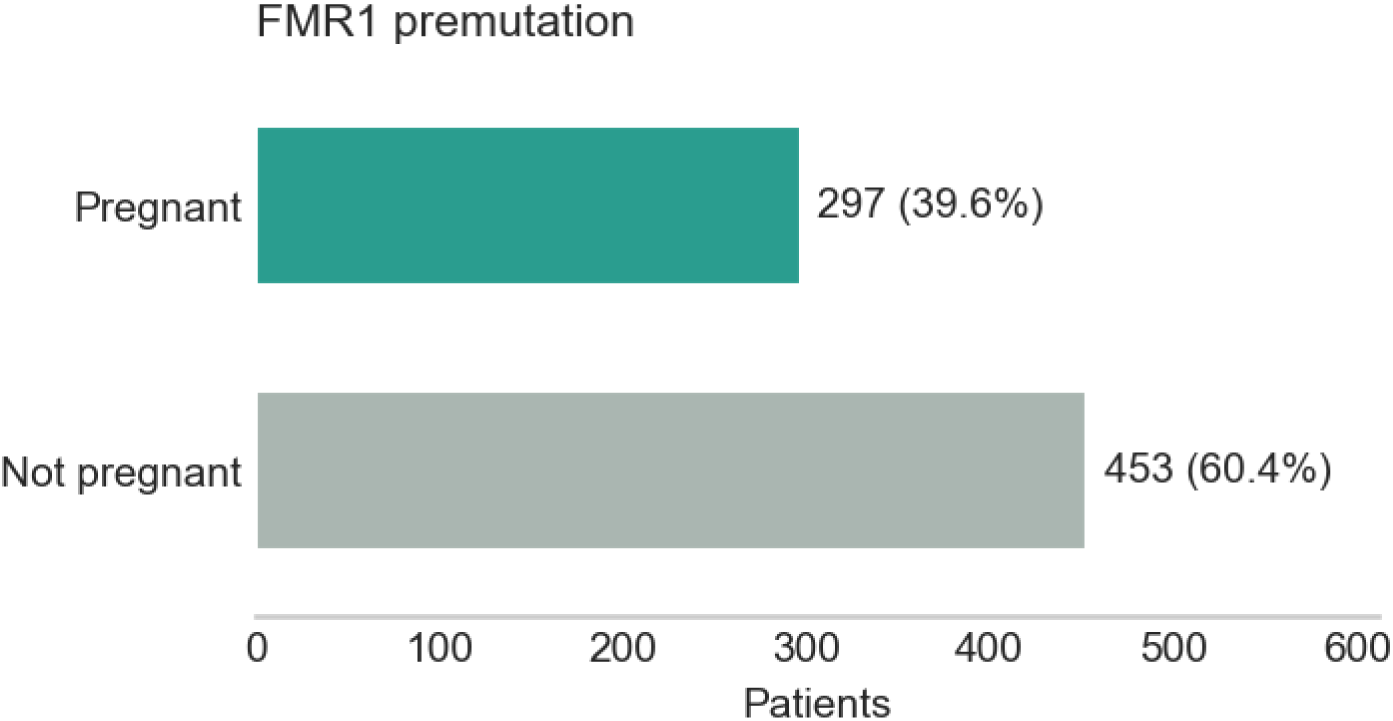
Documented pregnancy evidence in the confirmed *FMR1* premutation cohort. Bar chart of female patients (n = 739) with (297; 40.2%) and without (442; 59.8%) laboratory-confirmed pregnancy evidence, as defined in Methods.

**Figure 4:**
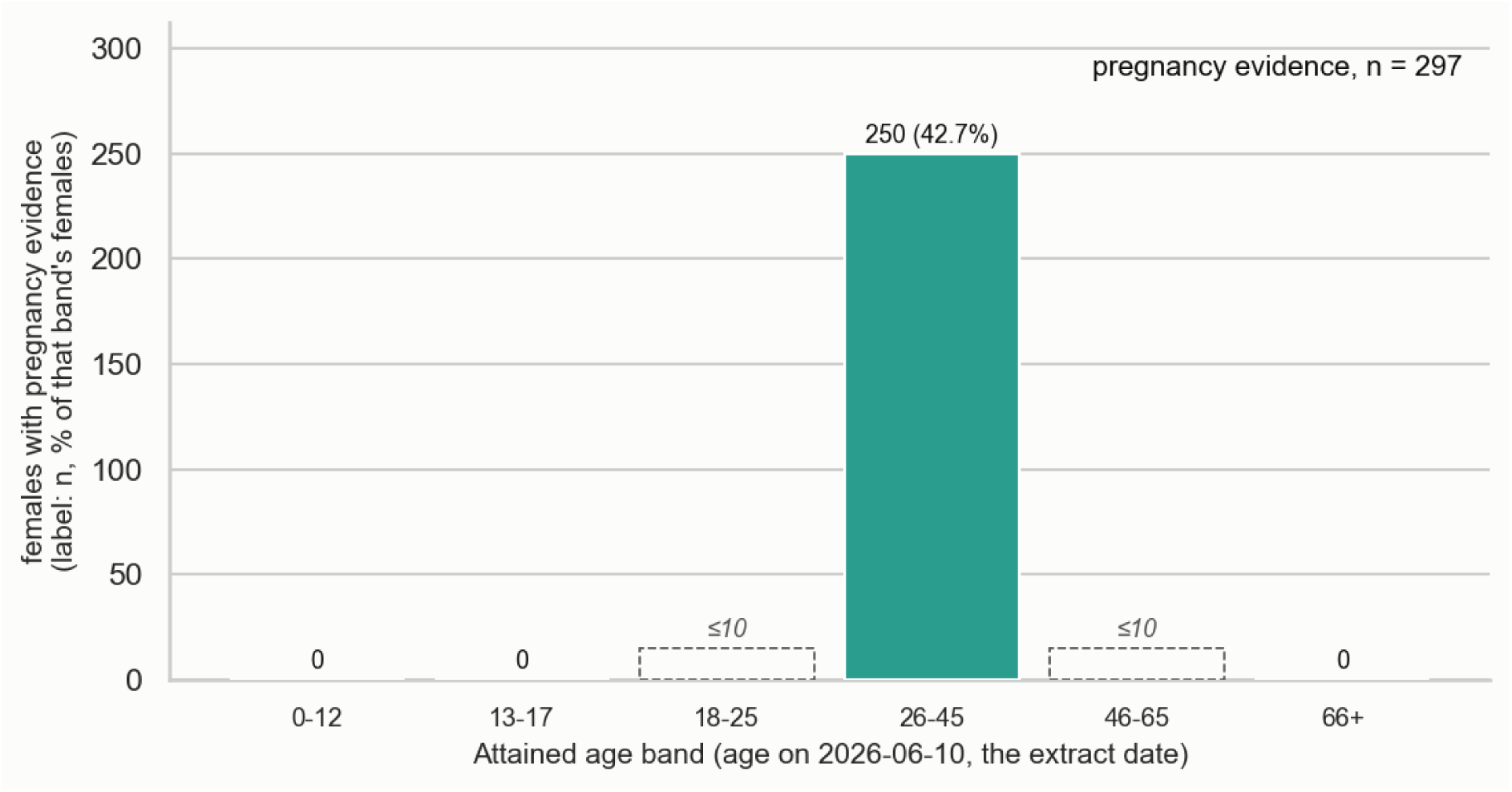
Age distribution of female patients with documented pregnancy evidence (n = 297). Bars show the number of females with pregnancy evidence in each attained-age band (age on the data-extraction date, 10 June 2026); labels give n and the proportion of all females in that band. Almost all pregnancy evidence falls in the 26–45-year band (250; 42.7% of females in that band); counts in the 18–25 and 46–65 bands are 1–10 and are shown as ≤10. The concentration in the reproductive age range is consistent with identification through prenatal carrier screening.

### Comorbidity Characterisation

Across the *FMR1* premutation cohort, endocrine/metabolic conditions (16.5%) and neuropsychiatric conditions within the FXAND domain (16.3%) were the most frequently documented phenotypic categories, followed by other conditions (8.8%) and reproductive/ovarian conditions within the FXPOI domain (8.5%). Pain/fatigue (2.5%), autoimmune/inflammatory conditions (2.0%), and neurological conditions within the FXTAS domain (1.7%) were documented less frequently. No comorbidities were identified in 61.9% of individuals in the cohort (Figure 5). Figure 6 illustrates the temporal relationship between first documentation of FXPAC-associated phenotypic domains and *FMR1* premutation carrier recognition, highlighting phenotypic domains documented prior to genetic recognition.

**Figure 5:**
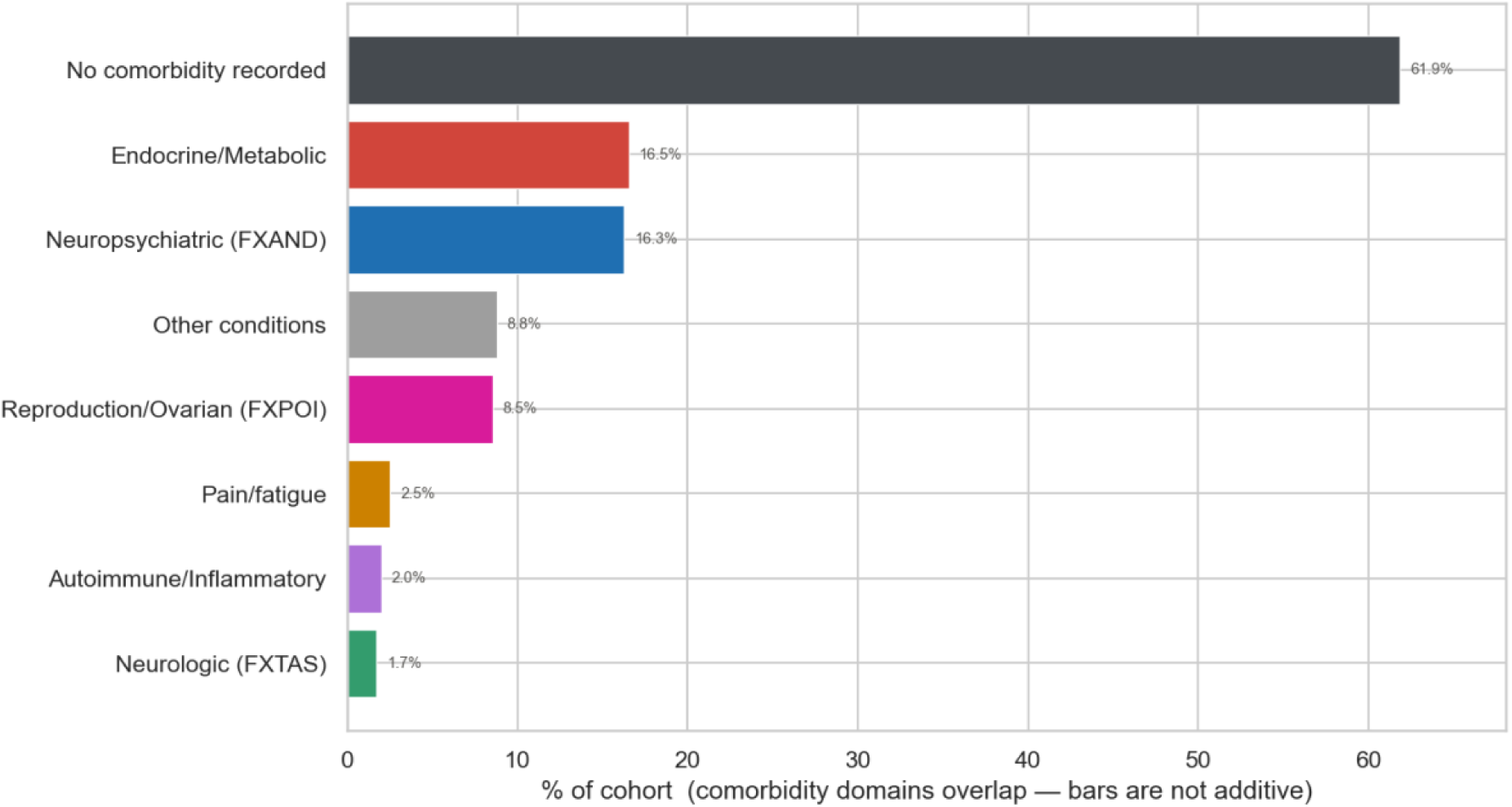
Phenotype domain prevalence in *FMR1* premutation carriers. Horizontal bar chart showing the prevalence of seven FXPAC-associated phenotypic domains (neuropsychiatric/FXAND, endocrine/metabolic, other, reproductive/FXPOI, pain/fatigue, neurological/FXTAS, autoimmune/inflammatory) among confirmed *FMR1* premutation carriers.

**Figure 6:**
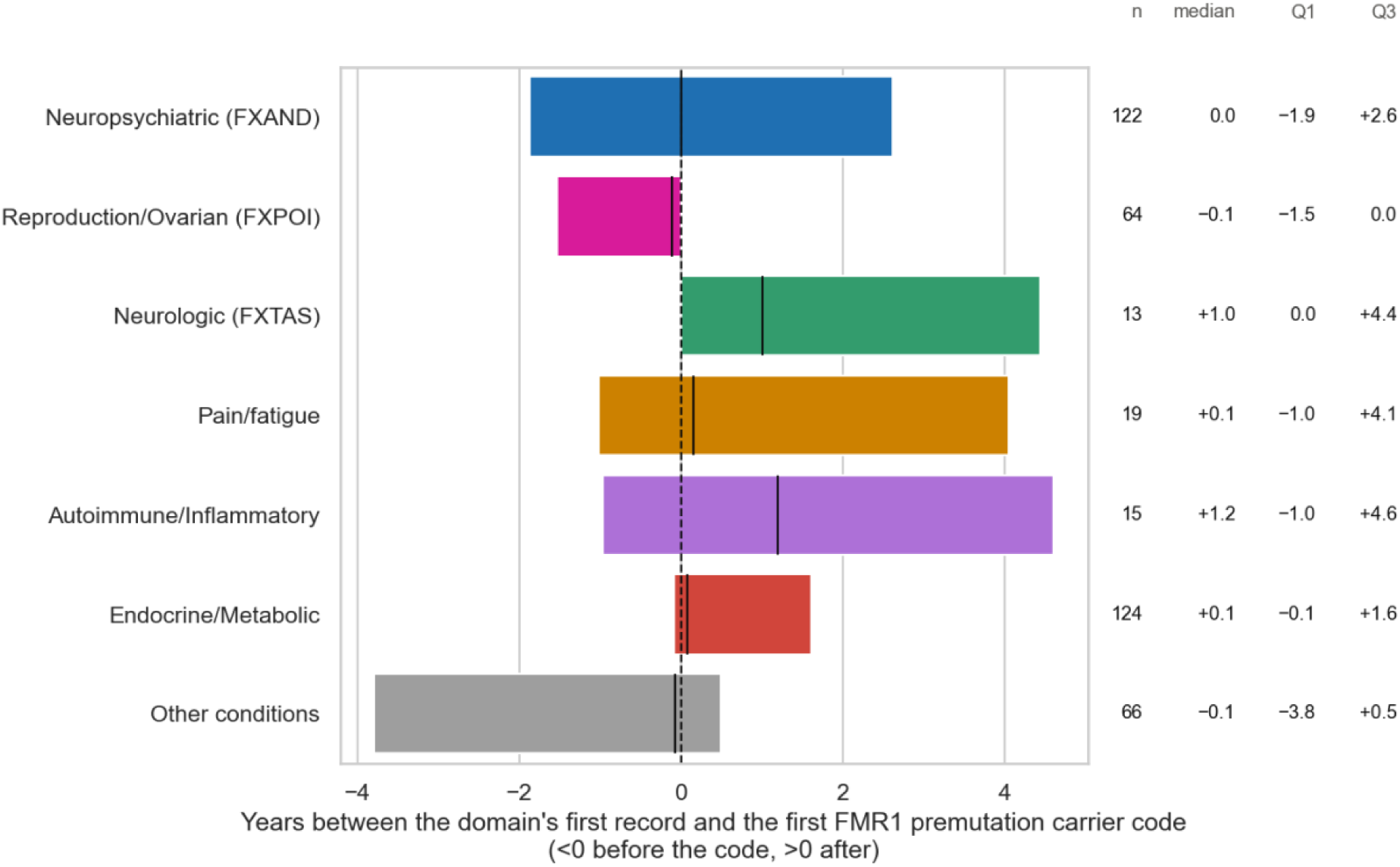
Timing of first documentation of each FXPAC-associated phenotypic domain relative to the first *FMR1* premutation carrier code. For each of the seven domains, the box spans the interquartile range of the lag in years between the earliest record in that domain and the carrier code, with the vertical line marking the median; whiskers are omitted. Negative values indicate documentation before the carrier code (dashed line at 0). The right-hand columns give the number of affected patients and the median, first and third quartiles in years. Intervals are documentation-to-documentation and do not represent biological onset.

### Diagnostic Trajectory Prior to Carrier Identification

Analysis of the timing of clinical diagnoses relative to the first recorded *FMR1* Premutation code showed that several key FXPAC-associated conditions were frequently documented in the EHR prior to *FMR1* Premutation identification. Table 2 summarises, for each of the seven phenotypic domains, the number of affected patients whose first record in that domain fell before, on, or after the index date, together with the median and interquartile range of the lag. Pre-index documentation was most frequent for reproductive/ovarian conditions (FXPOI), recorded before carrier identification in 38 of 64 affected patients (59.4%; median lag, −43 days; IQR, −561 to 0), and for the heterogeneous "other conditions" domain (36 of 66; 54.5%; median, −30 days; IQR, −1,386 to 178). Neuropsychiatric conditions (FXAND) preceded the carrier code in 56 of 122 patients (45.9%), with a median lag of 0 days but a widespread (IQR, −683 to 954 days), indicating that roughly a quarter of affected patients had a neuropsychiatric diagnosis more than 1.9 years before the carrier code. Endocrine/metabolic conditions were more often documented after identification (68 of 124; 54.8%) than before (42; 33.9%), with a median lag of +27 days. For the three smaller domains (FXTAS, n = 13; pain/fatigue, n = 19; autoimmune/inflammatory, n = 15) all before/same-day/after counts were below the disclosure threshold and are masked; their positive median lags (365, 53 and 435 days, respectively) suggest that these conditions were more commonly recorded after carrier identification. The distribution of diagnostic lags for each condition is shown in Figure 7 and Table 2.

**Figure 7:**
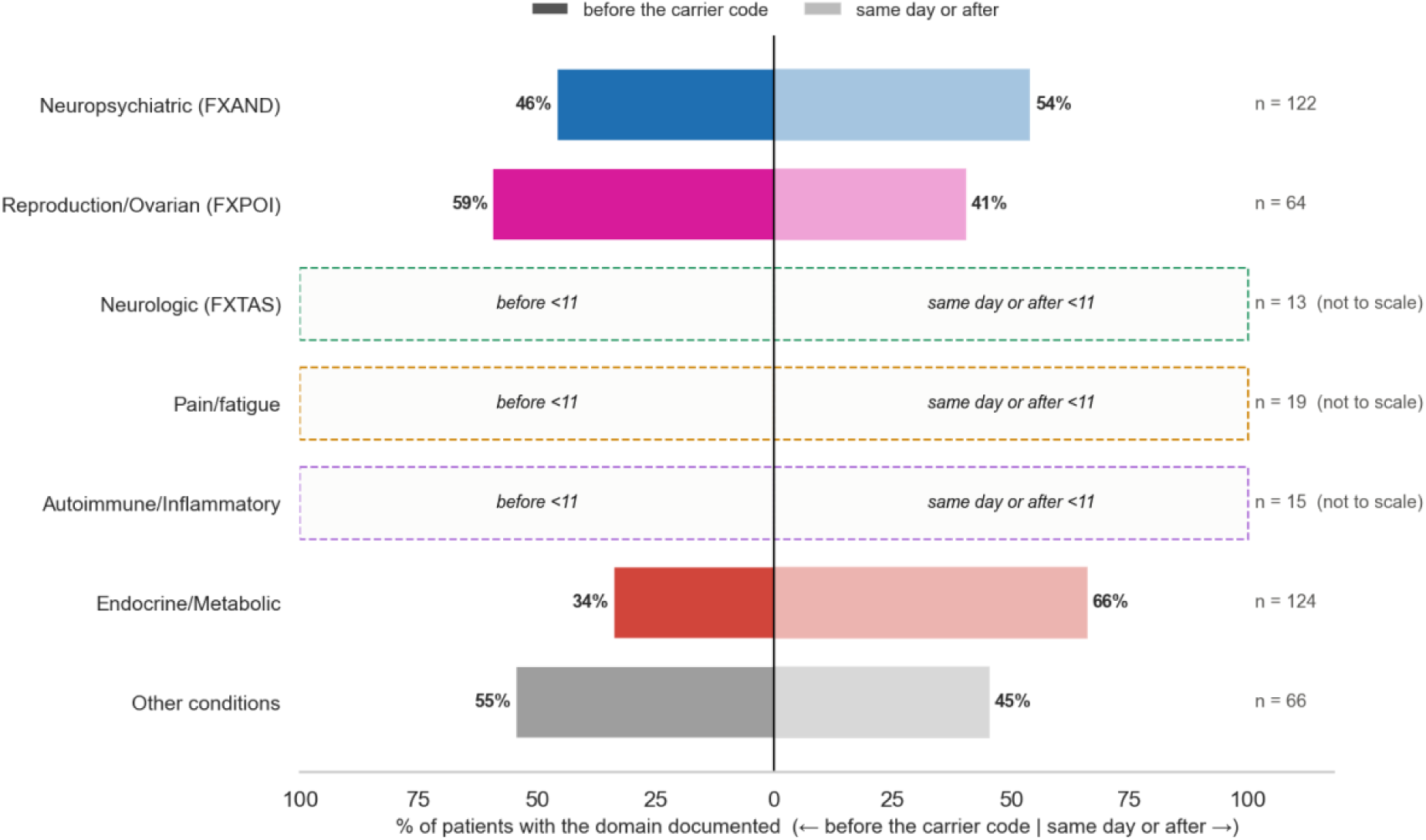
Diagnostic trajectory relative to the first *FMR1* premutation carrier code. Bar plot showing the percentage of population having recorded comorbidity diagnosis before and after recording the first *FMR1* premutation code for seven FXPAC-associated phenotypic domains; for domains in which either count is 1–10 (Neurologic/FXTAS, Pain/fatigue, Autoimmune/Inflammatory) the bars are drawn as dashed outlines and are not to scale. Negative values indicate that the condition was diagnosed before genetic identification, and positive values show that it was after genetic identification. Numbers show percentage of the population who had a diagnosis before or after genetic identification.

**Table 1:** Characteristics of the confirmed *FMR1* premutation cohort (n = 750): demographics, pregnancy evidence and prevalence of FXPAC-associated phenotypic domains.

| Characteristic | Denominator | *FMR1* Premutation | Evidence grade | References |
| --- | --- | --- | --- | --- |
| **Cohort** |  |  |  |  |
| **N patients, n** | patients in cohort | 750 |  |  |
| **Demographics** |  |  |  |  |
| **Age at index diagnosis (years), median [IQR]** | known age at index | 33.3 [30.0, 36.8] |  |  |
| **Female, n (%)** | patients in cohort | 739 (98.5) |  |  |
| **Any pregnancy evidence, n (%)** | females in cohort | 297 (40.2) |  |  |
| **Comorbidity (high-level group)** |  |  |  |  |
| **Neuropsychiatric (FXAND), n (%)** | patients in cohort | 122 (16.3) | Emerging (conflicting) | 5-7 |
| **Reproduction/Ovarian (FXPOI), n (%)** | patients in cohort | 64 (8.5) | Well-established | 4,7,9 |
| **Neurologic (FXTAS), n (%)** | patients in cohort | 13 (1.7) | Well-established | 18-20 |
| **Pain/fatigue, n (%)** | patients in cohort | 19 (2.5) | Emerging | 8,10,20-22 |
| **Autoimmune/Inflammatory, n (%)** | patients in cohort | 15 (2.0) | Emerging | 22 |
| **Endocrine/Metabolic, n (%)** | patients in cohort | 124 (16.5) | Emerging (thyroid only) | 20-23 |
| **Other conditions, n (%)** | patients in cohort | 66 (8.8) | - | 24 |

1. Continuous variables: median [IQR]; mean (SD) shown additionally only where the pooled distribution is roughly symmetric ( $|\text{skew}| \leq 0.5$ ).
2. Categorical variables: n (%); the denominator column names the base of each percentage. The pregnancy row uses the number of FEMALES in the cohort, not the cohort total, and counts a patient once if any evidence of pregnancy is recorded for her.
3. The table is descriptive; no significance testing is reported.

**Table 2:** Timing of FXPAC-associated diagnoses relative to first *FMR1* identification.

| Domain | n | Before | Same day | After | Lag, days: median [IQR] |
| --- | --- | --- | --- | --- | --- |
| **Neuropsychiatric (FXAND)** | 122 | 56 (45.9%) | <11 | <11 | 0 [-683, 954] |
| **Reproduction/Ovarian (FXPOI)** | 64 | 38 (59.4%) | 11 (17.2) | 15 (23.4) | -43 [-561, 0] |
| **Neurologic (FXTAS)** | 13 | <11 | <11 | <11 | 365 [0, 1623] |
| **Pain/fatigue** | 19 | <11 | <11 | <11 | 53 [-373, 1480] |
| **Autoimmune/Inflammatory** | 15 | <11 | <11 | <11 | 435 [-356, 1682] |
| **Endocrine/Metabolic** | 124 | 42 (33.9 %) | 14 (11.3) | 68 (54.8) | 27 [-30, 587] |
| **Other conditions** | 66 | 36 (54.5%) | <11 | <11 | -30 [-1386, 178] |

1. Lag in days = first recorded date of the condition minus the index date (first recorded carrier code); negative values mean the condition was documented before the carrier code. Same-day documentation is reported in its own column.
2. A patient is counted as affected in a domain when any condition in that domain is recorded; the domain's date is the earliest such record, so each patient contributes once per domain.
3. These are documentation-to-documentation intervals, not biological onset: both endpoints are coding dates, so the table describes when conditions enter the record relative to the carrier code, not causal ordering.
4. Domains overlap: a patient may appear in more than one, so rows are not mutually exclusive and do not sum to the cohort.
5. Percentages are of n affected in that domain. Disclosure control: counts of 1-10 are masked as '<11' and their percentages withheld.

## Discussion

In this large-scale EHR-based phenotyping study of *FMR1*-premutation carriers at a referral centre, we characterised the clinical phenotype of 750 patients with confirmed *FMR1* premutation status, which is, to our knowledge, the largest *FMR1* premutation cohort described to date within a single health system. The demographic profile of the confirmed *FMR1* premutation cohort, that is, predominantly women of reproductive age (median age at index 33.3 years), with two in five (40.2%) having laboratory-confirmed pregnancy evidence, is strongly consistent with ascertainment through routine prenatal carrier screening rather than clinical recognition of FXPAC. This distinction has important implications for interpretation, namely, that prevalence estimates from this cohort are more likely to reflect a screening-identified population than a clinically ascertained one. Future work using Natural Language Processing (NLP) on clinical notes and Generative AI (GenAI) may increase cohort size, substantially expanding the analytically confirmed cohort.

Our findings reveal a consistent and clinically meaningful comorbidity pattern already established in the spectrum of FXPAC, in the form of a substantial share of key *FMR1* premutation-associated diagnoses, including infertility, ADHD, depression, and anxiety etc. Most of them were often documented in the EHR before the *FMR1* premutation carrier status was formally recorded. This diagnostic gap underscores a critical and largely unaddressed challenge in the clinical management of *FMR1* premutation carriers: that the genetic underpinning of their symptoms frequently goes unrecognised long after those symptoms have presented. The findings have direct implications for how EHR data can be leveraged to reduce diagnostic delay, support earlier genetic referral, and ultimately improve outcomes for the affected population and underscore the central clinical need for an *FMR1* premutation cohort.

These patterns likely reflect multiple pathways through which *FMR1* premutation carriers are identified in the health system: clinical evaluation prompted by symptoms or associated conditions; reproductive carrier screening, often conducted as part of routine pregnancy care; and family cascade testing following identification of an affected relative. Patients presenting with FXPAC-associated symptoms may be managed for years under diagnostic labels without capturing underlying genetic aetiology and condition-specific treatment ^25–27^ . These findings align with prior qualitative evidence of diagnostic delay in FXTAS and FXPOI and provide population-level quantification of that delay in a real-world clinical cohort.

The combination of a substantial carrier prevalence ^2^, with the systematic diagnostic delays documented here positions the *FMR1* premutation as a public health issue of underappreciated magnitude. Most of the FXPAC identified in this cohort (anxiety, ADHD, infertility) appear frequently in the general population ^28–31^ and are routinely managed in primary and secondary care without necessarily prompting genetic investigation. Population-level EHR-based screening algorithms using precursor diagnosis patterns could, in principle, flag individuals for preventive management. Such approaches would need to balance sensitivity against specificity, given the non-specificity of many FXPAC-associated diagnoses. Nevertheless, the present findings provide an empirical basis for exploring whether EHR-derived phenotypic signatures could support earlier identification of at-risk individuals.

EHR-based carrier identification requires the acknowledgement of the ethical complexity of this approach. The majority of *FMR1* premutation carriers live full, healthy lives without ever developing FXTAS, FXPOI, or clinically significant FXAND. The penetrance of these conditions is incompletely predicted by CGG repeat size alone. EHR-based flagging of individuals as potential *FMR1* premutation carriers based on precursor diagnoses would, if implemented clinically, raise important questions. For example, who should be informed, under what circumstances, and by whom? Unsolicited carrier identification may impose a substantial psychological burden, particularly when the condition’s long-term trajectory is uncertain, and may have important implications for insurance, employment, and reproductive decision-making. While the research presented here is observational and retrospective, ethical considerations are essential for any translational application of these findings.

This study demonstrates the feasibility and value of applying OMOP-mapped EHR data to characterise rare genetic conditions at the population scale. The use of standardised concept codes enables reproducibility of the cohort definition at other institutions using the OMOP CDM, supporting future multi-site replication and external validation. Longitudinal EHR data enable analysis of diagnostic trajectories that is often not possible in traditional clinical studies or disease registries, which typically capture cross-sectional snapshots of the phenotype. The approach described here provides a scalable template for the systematic characterisation of other rare genetic conditions, particularly those with multi-system involvement and long diagnostic odysseys.

However, several limitations of this study must be acknowledged. First, *FMR1* premutation carrier status was primarily ascertained from EHR diagnostic codes. Unavailability of structured genetic tests weakens causal confirmation of *FMR1* premutation cohort. This illustrates a common limitation of observational EHR research: diagnostic coding may be incomplete or insufficiently specific. Further deep phenotyping using additional data modalities, particularly laboratory results and clinical notes, is therefore needed. In the future, we intend to apply Natural Language Processing (NLP) to clinical notes of patients to expand the *FMR1* premutation cohort. Furthermore, the ICD codes used for comorbidity ascertainment capture broad clinical categories: for example, the tremor code (R25.1) encompasses essential tremor and other etiologies beyond FXTAS, and anxiety codes do not distinguish FXAND-related from idiopathic presentations. These limitations are inherent to observational EHR research and are common to all such studies.

The present study establishes a foundation for several important extensions. Integration of CGG repeat count data, where available as structured results, would allow stratification of phenotypic burden by repeat size and more precise molecular characterisation of the cohort. We also expect to further sub-phenotype the *FMR1* premutation cohort into various FXPAC outcomes and create a prediction model to predict outcomes for *FMR1* premutation carriers in an effort to help mitigate the onset of various FXPAC conditions. Machine learning models trained on patterns of precursor diagnosis may ultimately support the development of EHR-based carrier detection algorithms. Finally, expanding this approach to the *FMR1* full mutation cohort at Mount Sinai will enable direct comparison of phenotypic burden and diagnostic trajectories across the *FMR1* expansion spectrum.

## Data availability (statement)

Data was made available to qualified researchers through appointment as adjunct faculty or visiting researchers at the Icahn School of Medicine at Mount Sinai and inclusion on the relevant IRB protocol, approved by the Icahn School of Medicine at Mount Sinai Institutional Review Board (STUDY-19-00951: HPIMS Data Science Protocol), with a waiver of informed consent for the secondary use of clinical data.

## Code availability

Will be shared on request.

## Acknowledgments

AIR·MS: This work is supported in part through the use of the research platform AI-Ready Mount Sinai (AIR·MS) and the expertise provided by the team at the Hasso Plattner Institute for Digital Health at Mount Sinai (HPIMS).

Minerva: This work was supported in part through the Minerva computational and data resources and staff expertise provided by Scientific Computing and Data at the Icahn School of Medicine at Mount Sinai and supported by the Clinical and Translational Science Awards (CTSA) grant UL1TR004419 from the National Centre for Advancing Translational Sciences.

## Author contributions

AR, EA and RL conceptualized and wrote the manuscript. AR, AA and JE performed the analysis. GN and LHW contributed and edited the manuscript. The final version of the manuscript was critically reviewed and approved by all authors.

## References

1 Movaghar, A. et al. Data-driven phenotype discovery of FMR1 premutation carriers in a population-based sample. Sci Adv 5, eaaw7195 (2019). 10.1126/sciadv.aaw7195

2 Seltzer, M. M. et al. Prevalence of CGG expansions of the FMR1 gene in a US population-based sample. Am J Med Genet B Neuropsychiatr Genet 159B, 589–597 (2012). 10.1002/ajmg.b.32065

3 Movaghar, A., Page, D., Brilliant, M. & Mailick, M. Prevalence of Underdiagnosed Fragile X Syndrome in 2 Health Systems. JAMA Netw Open 4, e2141516 (2021). 10.1001/jamanetworkopen.2021.41516

4 Allen, E. G. et al. Refining the risk for fragile X-associated primary ovarian insufficiency (FXPOI) by FMR1 CGG repeat size. Genet Med 23, 1648–1655 (2021). 10.1038/s41436-021-01177-y

5 Flavell, J., Franklin, C. & Nestor, P. J. A Systematic Review of Fragile X-Associated Neuropsychiatric Disorders. J Neuropsychiatry Clin Neurosci 35, 110–120 (2023). 10.1176/appi.neuropsych.21110282

6 Chubick, A., Wang, E., Au, C., Grody, W. W. & Ophoff, R. A. Large-Scale Whole Genome Sequence Analysis of >22,000 Subjects Provides no Evidence of FMR1 Premutation Allele Involvement in Autism Spectrum Disorder. Genes (Basel) 14 (2023). 10.3390/genes14081518

7 Klausner, L. et al. No association between FMR1 premutation and either ADHD or anxiety in 53,707 women undergoing genetic testing for family planning purposes. Genet Med 27, 101428 (2025). 10.1016/j.gim.2025.101428

8 Pinheiro, M. et al. Advances in exploring the association between FMR1 premutation and fibromyalgia: a pilot study with a more effective sample definition. Clinics (Sao Paulo) 80, 100758 (2025). 10.1016/j.clinsp.2025.100758

9 Morbey, E. J. et al. Large-scale analysis of FMR1 CGG repeat length and risk of premature ovarian insufficiency in over 92 000 women. Hum Reprod 41, 998–1007 (2026). 10.1093/humrep/deag061

10 Protic, D. et al. Chronic pain, fatigue, and emotional distress in female FMR1 premutation carriers. Front Mol Neurosci 19, 1741854 (2026). 10.3389/fnmol.2026.1741854

11 Montanaro, F. A. M. et al. Psychiatric and Cognitive Features in Italian Women With the FMR1 Premutation: A Comprehensive Assessment Using SCID-5 and Standardized Cognitive Measures. Am J Med Genet B Neuropsychiatr Genet 201, 406–419 (2026). 10.1002/ajmg.b.70014

12 Lozano, R., Rosero, C. A. & Hagerman, R. J. Fragile X spectrum disorders. Intractable & Rare Diseases Research 3, 134–146 (2014). 10.5582/irdr.2014.01022

13 Lozano, R., Azarang, A., Wilaisakditipakorn, T. & Hagerman, R. J. Fragile X syndrome: A review of clinical management. Intractable Rare Dis Res 5, 145–157 (2016). 10.5582/irdr.2016.01048

14. Lozano, R., et al. Observable Symptoms of Anxiety in Individuals with Fragile X Syndrome: Parent and Caregiver Perspectives. Genes (Basel) 13 (2022). 10.3390/genes13091660

15 Tassone, F. et al. Insight and Recommendations for Fragile X-Premutation-Associated Conditions from the Fifth International Conference on FMR1 Premutation. Cells 12 (2023). 10.3390/cells12182330

16 Hagerman, R. J. & Hagerman, P. J. The Spectrum of Fragile X Disorders. New England Journal of Medicine 393, 281–288 (2025). doi:10.1056/NEJMra2300487

17 Guerrero, P. et al. The AIR.MS data platform for artificial intelligence in healthcare. JAMIA Open 8, ooaf145 (2025). 10.1093/jamiaopen/ooaf145

18 Jacquemont, S. et al. Fragile X premutation tremor/ataxia syndrome: molecular, clinical, and neuroimaging correlates. Am J Hum Genet 72, 869–878 (2003). 10.1086/374321

19 Jacquemont, S. et al. Penetrance of the fragile X-associated tremor/ataxia syndrome in a premutation carrier population. JAMA 291, 460–469 (2004). 10.1001/jama.291.4.460

20 Rodriguez-Revenga, L. et al. Penetrance of FMR1 premutation associated pathologies in fragile X syndrome families. Eur J Hum Genet 17, 1359–1362 (2009). 10.1038/ejhg.2009.51

21 Coffee, B. et al. Mosaic FMR1 deletion causes fragile X syndrome and can lead to molecular misdiagnosis: a case report and review of the literature. Am J Med Genet A 146A, 1358–1367 (2008). 10.1002/ajmg.a.32261

22 Winarni, T. I. et al. Immune-mediated disorders among women carriers of fragile X premutation alleles. Am J Med Genet A 158A, 2473–2481 (2012). 10.1002/ajmg.a.35569

23 Lozano, R. et al. Aging in Fragile X Premutation Carriers. Cerebellum 15, 587–594 (2016). 10.1007/s12311-016-0805-x

24 Au, J. et al. Prevalence and risk of migraine headaches in adult fragile X premutation carriers. Clin Genet 84, 546–551 (2013). 10.1111/cge.12109

25 Leehey, M. A., Hall, D.A., Liu, Y., Hagerman, R.J. . *Clinical Neurological Phenotype of FXTAS.*, (Springer, Cham., 2016).

26 Hall, D. A. & Hagerman, R. J. Fragile X-Associated Tremor/Ataxia Syndrome: Unmet Needs and a Path for the Future. Front Genet 9, 100 (2018). 10.3389/fgene.2018.00100

27 Hipp, H. S., Charen, K. H., Spencer, J. B., Allen, E. G. & Sherman, S. L. Reproductive and gynecologic care of women with fragile X primary ovarian insufficiency (FXPOI). Menopause 23, 993–999 (2016). 10.1097/GME.0000000000000658

28 Kroenke, K., Spitzer, R. L., Williams, J. B., Monahan, P. O. & Lowe, B. Anxiety disorders in primary care: prevalence, impairment, comorbidity, and detection. Ann Intern Med 146, 317–325 (2007). 10.7326/0003-4819-146-5-200703060-00004

29 Song, P. et al. The prevalence of adult attention-deficit hyperactivity disorder: A global systematic review and meta-analysis. J Glob Health 11, 04009 (2021). 10.7189/jogh.11.04009

30 Cortese, S. et al. Incidence, prevalence, and global burden of attention-deficit/hyperactivity disorder from 1990 to 2021 across 204 countries in individuals under age 20: data, with critical appraisal, from the 2021 Global Burden of Disease study. Mol Psychiatry (2026). 10.1038/s41380-026-03683-4

31 Fernandez, R. C. et al. Trends in clinical encounters and management for infertility among women attending Australian general practice: a national longitudinal study using MedicineInsight, 2011 to 2021. BMJ Open **15**, e085149 (2025). 10.1136/bmjopen-2024-085149

